# Rising Through the Pandemic: A scoping review of quality improvement in public health during the COVID-19 pandemic

**DOI:** 10.1101/2021.06.08.21258002

**Authors:** X. Cindy Yin, Michelle Pang, Madelyn Law, Fiona Guerra, Tracey O’Sullivan, Rachel E. Laxer, Brian Schwartz, Yasmin Khan

## Abstract

**Background:** The COVID-19 pandemic generated a growing interest in and need for evidence-based tools to facilitate the implementation of emergency management strategies within public health practice. Quality improvement (QI) has been identified as a key framework and philosophy to guide organizational emergency response efforts; however, the nature and extent to which it has been used in public health settings during the COVID-19 pandemic remains unclear.

**Methods:** We conducted a scoping review of literature published January 2020 – February 2021 and focused on the topic of QI at public health agencies during the COVID-19 pandemic. The search was conducted on four bibliographic databases, in addition to a supplementary grey literature search using custom Google search engines and targeted website search methods. Of the 1,878 peer-reviewed articles assessed, 15 records met the inclusion criteria. An additional 11 relevant records were identified during the grey literature search, for a total of 26 records included in the scoping review.

**Results:** Records were organized into five topics: 1) collaborative problem solving and analysis with stakeholders; 2) supporting learning and capacity building in QI; 3) learning from past emergencies; 4) implementing QI methods during COVID-19; and 5) evaluating performance using frameworks/indicators.

**Conclusions:** The literature indicates that QI-oriented activities are occurring at the organizational and program levels to enhance COVID-19 response. To optimize the benefits that QI approaches and methodologies may offer, it is important for public health agencies to focus on both widespread integration of QI as part of an organization’s management philosophy and culture, as well as project level activities at all stages of the emergency management cycle.

## 1. Background

The Coronavirus Disease 2019 (COVID-19) pandemic generated a growing interest in and need for evidence-based tools and techniques to facilitate the implementation of public health emergency management (PHEM) strategies. Quality improvement (QI) is one such approach, and is defined as “the use of deliberate and defined methods in continuous efforts to achieve measurable improvements in the efficiency, effectiveness, performance, accountability, outcomes, and other indicators of quality in services or processes,.”^1^ QI – in a more broad sense – is used as a management philosophy to guide PHEM during large-scale infectious disease emergencies; however, its relevance in the complexity of the COVID-19 pandemic warrants exploration.

Emergencies are multifaceted, requiring strong and wide-scale coordination and collaboration across sectors, community partners and within public health agencies. Emergencies also require adaptive and efficient processes to support an effective response. The nature of response efforts render the use of QI approaches useful, given the focus is on adapting programs, services and practices in real-time, as well as the measurement and improvement of system performance on an ongoing basis. Moreover, response requires flexibility and continuous adaptation to the changing aspects of a pandemic, which aligns with the methods and approaches central to QI.

While QI is a well-established field of study and practice in clinical health care settings, the state of QI in public health – where quality-driven programs, services, policies and research for improved health outcomes and conditions^2^ – is still emerging. Previous research has sought to clarify the role of QI in public health^3^ and public health emergency preparedness.^4,5^ In 2007, Seid and colleagues described a “preparedness production system”, whereby public health agencies engage in routine, systematic activities to bolster capability-building and ongoing surveillance/detection before an emergency occurs.^4^ This ongoing work may help to prepare public health agencies and stakeholders for an optimal response, leading to improved outcome indicators such as reduced morbidity, mortality, and social disruption after the emergency event.^4^

In practice, the understanding and application of QI in public health settings ranges from individual small-scale projects implemented at a programmatic level, to agency-wide implementation of QI frameworks as part of an organization’s culture.^1,6^ Both of these elements are essential to support improvement efforts. Having a QI-oriented management philosophy and supporting structures that allow staff to engage in associated methodology and individual project-level QI activities ensures alignment and enhancement of existing practice. Specifically, on the individual team or project level, formal QI tools and techniques such as Plan-Do-Study-Act (PDSA) cycles (Model for Improvement), process mapping, Strengths/Weaknesses/Opportunities/Threats (SWOT) analysis, In/intra-Action-Reviews (IAR), and After-Action-Reviews (AAR) offer structured ways for teams to integrate improvement principles into established processes. At the organizational level, additional frameworks and methods exist for broader implementation of QI throughout the organization. These include Lean enterprise, Six Sigma, continuous quality improvement, and other management frameworks and principles commonly seen in health care and other disciplines (e.g., engineering, manufacturing) where the integration of QI concepts in organizational processes and culture are more commonplace.

The COVID-19 pandemic elicited renewed interest in the capability and capacity of public health systems to respond to infectious disease emergencies. Before and during emergencies there are diverse opportunities to integrate QI in operations in order to ensure efficiency and effectiveness of core response activities (e.g., surveillance, case and contact management, vaccine distribution) and emergency management (e.g., implementation of emergency response plans, incident management structure activation) functions. Previous evidence syntheses explored how QI is operationalized in public health,^3,7,8^ yet information about the nature and extent to which QI methods, tools, and techniques have been implemented in PHEM settings during the COVID-19 pandemic is still emerging. The objective of this scoping review is to explore literature on applications of QI at both the organization-wide and project level at public health agencies during the pandemic (i.e., both the broader supporting structures and strategies are considered together with the implementation of QI tools, methods and specific individual projects). Information on QI experiences across different contexts and jurisdictions can inform the development and mobilization of PHEM strategies to enhance response to and recovery from the COVID-19 pandemic and future infectious disease emergencies.

## 2. Methods

To achieve the research objective, scoping review methodology was employed. Scoping reviews are a type of knowledge synthesis which aims to map existing literature on a new, complex or heterogeneous topic of interest with respect to its volume, nature and characteristics.^9^ Scoping reviews are commonly conducted to understand the state of the literature on a novel or emerging topic and to identify research gaps in the existing literature, and as such, it is a good option for exploring QI in the context of the COVID-19 pandemic.^9^ The literature search was conducted by a research team at Public Health Ontario (PHO), a provincial public health agency located in Toronto, Canada. The team consisted of individuals with training and expertise in public health science (QI, emergency management, infectious disease outbreaks) and research synthesis. PHO Library Information Specialists were consulted during search strategy development and involved in the article retrieval process.

### 2.1. Objective, research question, and scope

The objective of this review was to explore the current evidence base related to applications of QI at public health agencies during the COVID-19 pandemic, and was guided by two questions: “How have public health agencies used QI during the COVID-19 pandemic?” and “How can QI be used to support public health emergency management?” This review only includes QI initiatives undertaken by public health agencies (and other relevant PHEM settings) as well as QI resources applicable to these settings. Information on QI initiatives undertaken in patient/clinical care settings (e.g., primary care, emergency department) were out of scope. Studies and grey literature on QI initiatives related to clinical treatment for COVID-19, protective measures (e.g., distribution of personal protective equipment) and technical studies were also out of scope. Finally, this review did not examine the effectiveness of the QI initiatives or resources.

### 2.2. Data sources and search strategy

A search of both peer-reviewed and grey literature was conducted between January 2020 and February 2021. Four databases were selected to be comprehensive and inclusive of literature in the biomedical, public health, health science, and global health disciplines: MEDLINE, Ovid Embase, Ovid Global Health, and Scopus. Search strings used for the query were developed by PHO Library Information Specialists (see **Appendix A – Search Strings**) based on key terms deemed relevant to the topic by the research team, including (but not limited to): public health; quality improvement; novel coronavirus/SARS-CoV-2; COVID-19; health emergencies; and emergency management. The supplementary grey literature search was conducted by applying search strings to custom Google search engines tailored to generate results from relevant public health agency websites in Ontario, other provinces in Canada, the United States (US), and other international countries. Two reviewers (CY, MP) assessed the first 100 results from the Google search engines results and also conducted a targeted search of selected health QI agency webpages for relevant resources.

### 2.3. Eligibility criteria

Peer-reviewed articles were eligible for inclusion based on the following criteria: (1) takes place in a public health-related setting at any level (local/regional, national, international); (2) describes emergency management at any stage (mitigation, preparedness, response, recovery); (3) describes study objectives and/or methods based on QI; (4) uses an established QI approach, tool or technique (e.g., PSDA Cycles/Model for Improvement, SWOT analysis, root cause analysis), or uses qualitative/quantitative indicators and/or metrics to measure performance; and (5) describes implemented, supported or mandated actions. Records were considered ineligible if they were: basic research; epidemiological/clinical studies; medical/patient care research; clinical guidelines/best practices; and/or commentary/editorial/opinion pieces. In addition, records that described QI in non-human elements (e.g., methods for optimizing laboratory quality and safety, Electronic Medical Records updates) were excluded. Only English language articles focused on Organisation for Economic Co-operation and Development (OECD) countries published and between January 2020 and February 2021 were included in the search. The language restriction was placed due to limited resources for translation, while the location restriction was placed to capture publications from jurisdictions with similar or comparable public health system structures and contexts to Canada. The date restriction was placed to capture results with relevance to or discussion related to the COVID-19 pandemic.

For the supplementary grey literature search, English-language records related to OECD countries and published during the above date range were eligible for inclusion based on the following criteria: (1) published by a governmental health agency at any level (local/regional, national, international); (2) describes tools, techniques and/or resources for public health or related PHEM settings; and (3) describes tools, techniques and/or resources developed or using QI methodology.

### 2.4. Data screening and reference management

The peer-reviewed literature screening process involved two levels. For the first level of screening, titles and abstracts were reviewed by two independent reviewers (CY, FG). The second level of screening involved a full-text review by two reviewers (CY, MP) with any conflicting decisions resolved with by a third reviewer (FG). All screening, de-duplication, and reference management was completed using the systematic review software, Covidence.^10^ For the supplementary grey literature search, all relevant records were entered into a spreadsheet for tracking and processing. The final pool of peer-reviewed articles and grey literature records was reviewed and approved by four research team members (CY, MP, FG, YK).

### 2.5. Data extraction, summary, and synthesis

Data extraction was completed by two reviewers (CY, MP) based on the descriptive (i.e., year of publication, type of emergency, country, setting) and methodological (i.e., objective, methodology, data sources) characteristics of each record, in addition to key findings. Furthermore, each record was assessed for its QI relevance and emergency management cycle stage(s) discussed (i.e., preparedness, response, recovery, mitigation). The records were then grouped into overarching topics conceived by the research team, representing the nature in which QI has been operationalized during the COVID-19 pandemic.

## 3. Results

The peer-reviewed literature search returned a total of 1,878 records. After de-duplication and the first level of title and abstract review 74 were approved for a second level screening, of which 15 were eligible after screening based on the inclusion criteria (see **Figure 1 – PRISMA Flow Diagram for Peer-reviewed Literature Records)**. An additional 11 records were found during the supplementary grey literature search, for a total of 26 records. In total, the scoping review included 26 relevant records (see **Table 1 – Descriptive Summary of Records**).

**Figure 1.**
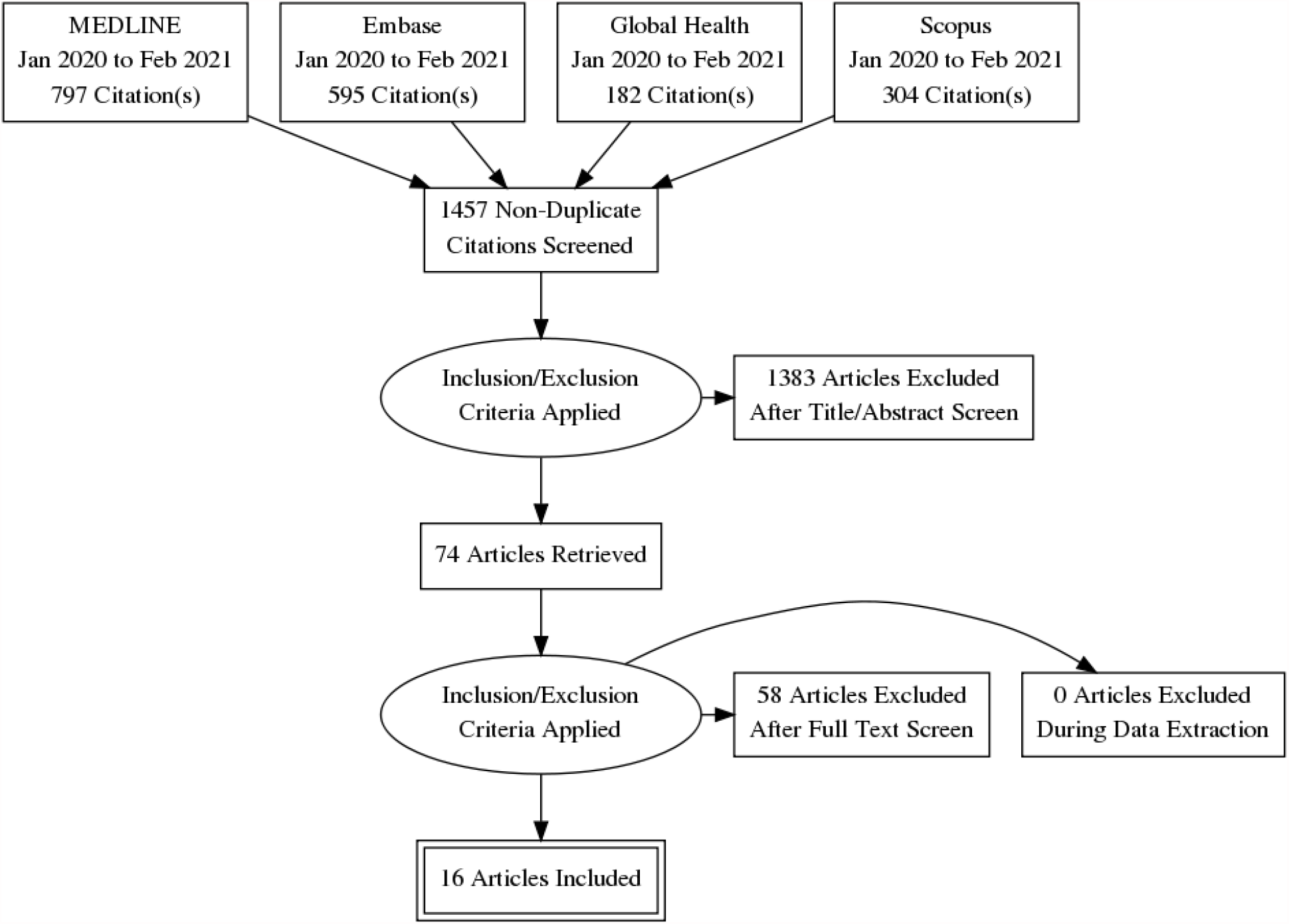
PRISMA Flow Diagram for Peer-reviewed Literature Records.

**Table 1.** Descriptive Summary of Records.

| Author/Organization: | Type: | Jurisdiction: | Setting: | QI Element: | Topics: |
| --- | --- | --- | --- | --- | --- |
| Advancing Quality Alliance & National Health Service<sup>41</sup> | Resource | UK | Health system | QI education | Learning and capacity building |
| Agency for Healthcare Research and Quality<sup>15</sup> | Resource | US | Health system | Learning community | Learning and capacity building |
| Aragon et al.<sup>12</sup> | Article | US (California) | PHU – Local/Regional | Framework<br>Root cause analysis<br>Checklist<br>PDSA | Collaborative problem solving and analysis with stakeholders<br>Learning and capacity building |
| Bacci et al.<sup>11</sup> | Article | US (Washington) | PHU – Local/Regional | Stakeholder engagement | Collaborative problem solving and analysis with stakeholders |
| Boyce et al.<sup>32</sup> | Article | US | PHU – Local/Regional | Framework | Use of framework/indicators |
| Centers for Disease Control and Prevention<sup>19</sup> | Program | US | PHU – Local/Regional | Training program | Learning and capacity building |
| Council of Ontario Medical Officers of Health<sup>17</sup> | Report | Canada (Ontario) | PHU – Local/Regional | Evaluation<br>Assessment (lessons learned) | Learning and capacity building |
| Curtis et al.<sup>29</sup> | Article | Australia | PHU – Local/Regional | Evaluation (CDC Framework) | Use of framework/indicators |
| First Nations Health Authority<sup>20</sup> | Program | Canada (BC) | Health system | Accreditation program | Learning and capacity building |
| Flynn et al.<sup>27</sup> | Article | US (Philadelphia) | Health care | Lessons learned<br>Process mapping | Use of framework/indicators<br>Learning and capacity building |
| Government of British Columbia<sup>42</sup> | Resource | Canada (BC) | Health system | After action review | Learning and capacity building<br>Collaborative problem solving and analysis with stakeholders<br>Learning from past emergencies |
| European Centre for Disease Prevention and Control<sup>22,23</sup> | Resource, Report | Europe | PHU – National | In action review<br>After action review<br>1 day in action review (condensed) | Learning and capacity building<br>Collaborative problem solving and analysis with stakeholders<br>Learning from past emergencies<br>QI methods during COVID-19 |
| Hamilton et al.<sup>43</sup> | Article | US | PHU – National | Assessment (challenges, recommendations) | Collaborative problem solving and analysis with stakeholders |
| Hunger et al.<sup>13</sup> | Article | Europe (Germany, Netherlands) | Health care | Checklist development | Collaborative problem solving and analysis with stakeholders |
| Kandel et al.<sup>31</sup> | Article | Multiple | PHU – International | Assessment | Use of framework/indicators<br>Learning and capacity building |
| Marshall et al.<sup>25</sup> | Article | US (Florida) | PHU – Local/Regional | Evaluation (WHO Health Systems Framework) | Use of framework/indicators<br>Learning from past emergencies |
| Mehta et al.<sup>44</sup> | Article | UK | Health care | Evaluation | Learning and capacity building<br>Collaborative problem solving and analysis with stakeholders<br>QI methods during COVID-19 |
| National Health Service<sup>28</sup> | Strategy | UK | Health system | Framework | Use of framework/indicators |
| Neogi et al.<sup>30</sup> | Article | Multiple | Health system | Evaluation (WHO Health Systems Framework) | Use of framework/indicators |
| Parker<sup>24</sup> | Article | Multiple | Health system | After action review | QI methods during COVID-19<br>Learning from past emergencies |
| RAND Europe<sup>45</sup> | Report | UK | PHU – National | Evaluation | Learning and capacity building |
| Ruebush et al.<sup>18</sup> | Article | USA | PHU – Local/Regional | Assessment (challenges, lessons learned) | Learning and capacity building<br>Collaborative problem solving and analysis with stakeholders |
| Toney et al.<sup>46</sup> | Article | USA | PHU – Laboratory | Assessment (challenges, recommendations) | QI methods during COVID-19<br>Learning and capacity building |
| Torri et al.<sup>14</sup> | Article | Europe (Italy) | PHU – Local/Regional | SWOT analysis | Collaborative problem solving and analysis with stakeholders<br>Learning and capacity building |
| World Health Organization<sup>21</sup> | Resource, Report | Multiple | PHU – International | Intra action review | QI methods during COVID-19<br>Learning from past emergencies<br>Collaborative problem solving and analysis with stakeholders<br>Learning and capacity building |

Records were identified from the following jurisdictions: Australia (n=1), Canada (n=3), Europe (multiple/unspecified country; [n=3]), Italy (n=1), United Kingdom (UK; [n=3]), US (n=11), and multiple/unspecified jurisdiction (n=4). Records were related to health care (n=3), laboratory (n=1), public health (local/regional level (n=9), national level (n=4), international/multi-jurisdictional (n=1) and health systems settings (n=8). Moreover, the records identified in this review discussed preparedness (n=2), preparedness/response (n=4), response (n=12), mitigation/preparedness/response (n=1), and all four stages of the emergency management cycle (n=1). This review did not find any records related solely to the recovery stage of the emergency management cycle. In addition, several records were related not to emergency management, but rather, overall health system improvement (n=6). (See **Figure 2 – Records by Emergency Management Cycle Phase**).

**Figure 2.**
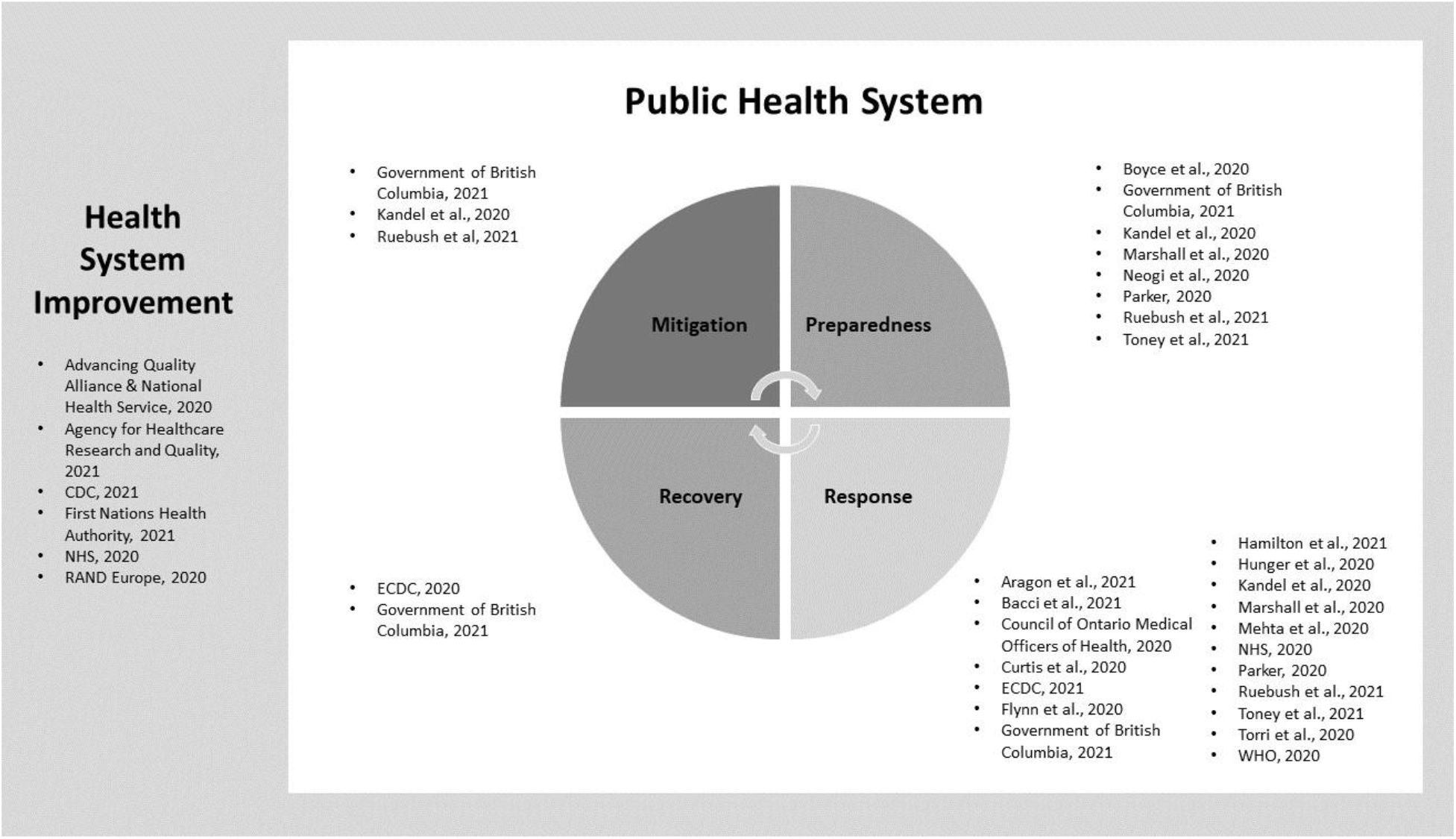
Records by Emergency Management Cycle Phase.

A variety of QI methods (e.g., learning communities, stakeholder engagement), tools and techniques (e.g., frameworks, performance indicators) and other improvement-related assessments (e.g., lessons learned, identifying challenges and opportunities) were discussed. Notably, the majority of grey literature records did not describe assessment of QI initiatives, but rather, provided resources developed for use in public health settings and were not specific to COVID-19.

### 3.1. Findings as organized into categories

The records identified during the review were organized based on the following five overarching topics, and reflect both organization-wide QI activities and project-based QI activities. These topics included: 1) collaborative problem solving and analysis with stakeholders; 2) supporting learning and capacity building in QI; 3) learning from past emergencies; 4) implementing QI methods during COVID-19; and 5) evaluating performance using frameworks/indicators. The topic identified were not mutually exclusive, as some studies discussed more than one area.

#### 3.1.1. Collaborative problem-solving and analysis with stakeholders

Ten records in this review described initiatives to facilitate collaborative problem-solving and analysis with stakeholders to bolster public health emergency response and improve response efforts. Several records highlighted interdependent efforts in working towards a shared goal of emergency preparedness and response. For example, a group from Washington State^11^ used a simulated pandemic influenza event to facilitate discussion exercises that identified strengths and opportunities between community pharmacy organizations, emergency preparedness officials from the local and state health departments, representatives of the state pharmacy association, and faculty from a school of pharmacy. The exercise was evaluated to validate strengths and improve capacity for participating organizations, with the expectation that every simulation would result in multiple findings and areas for improvement.^11^ Similarly, Aragon et al.^12^ showed that improving decision intelligence through QI methods and empirical evidence was a key element in building consensus and managing conflict across 13 Bay Area jurisdictions during the early response to COVID-19.

Research from Europe demonstrated that stakeholder engagement in real-time during the COVID-19 response helped to identify and leverage areas of shared knowledge regarding efficient collaboration, improved teamwork based on mutual respect, thus contributing to the development of innovative decision-making methods tailored to the needs of an inter-professional and multi-disciplinary COVID-19 response. Hunger et al.^13^ identified lessons learned from training, teaching and continuous feedback rounds to develop tailored training and methods to improve inter-professional collaboration and collaboration in mobile COVID-19 response teams. After the first wave of COVID-19 in Italy, Torri et al.^14^ conducted a SWOT analysis of the response strategies implemented by the Italian Department of Prevention and carried out by frontline health workers to examine which factors promoted or hindered their local response to COVID-19. The authors noted that it was crucial for decision-makers to understand the pandemic based on the local public health context, especially for individuals who work at in clinical health care settings where activity continuously changes throughout different phases of the pandemic.^14^ The article described complex processes which helped public health agencies and public health officials improve their decision-making and strategic planning, including a root cause analysis of the issues at hand.^14^ This use of root cause analysis aligns with the first phase of a QI approach to address changes to practice.

The importance of stakeholder participation in the collaborative decision-making response is increasingly being recognized as a process to improve pandemic response. The Agency for Healthcare Research and Quality (AHRQ)’s^15^ ACTS COVID-19 Evidence to Guidance to Action Collaborative program was found to exemplify stakeholder participation. In this program, participating organizations improve their work processes and results through stakeholder input. By collaborating with over 300 diverse entities, the AHRQ are developing a stakeholder-driven knowledge ecosystem that supports evidence to guidance to action to data and back^16^; an approach that is highly consistent with QI philosphy. Additionally, a report from the Council of Medical Officers of Health of Ontario^17^ highlighted a collaboration by local public health stakeholders to reflect on lessons learned and identify opportunities to improve the province of Ontario’s COVID-19 response. The use of evaluation and QI methods such as reviews, interviews and surveys helped local public health agencies maintain strong, collaborative relationship as they coordinated their responses to successive waves of COVID-19.

Collaborative problem-solving and analysis with stakeholders also emerged as a key topic in this scoping review. The response to the COVID-19 pandemic involves multiple levels of decision-making, often requiring real-time input from several organizations and levels of government in order to inform local-level improvements for response planning and action. It is important to note that this scoping focused on OECD countries, which generally have significant public health infrastructure consisting of public health agencies, health care organizations, and other entities with emergency management responsibilities. Given the multi-stakeholder context in complex emergencies, fascilitating stakeholder engagement strategies through an QI lens allows for reflection, evaluation and implementation that ultimately contributes to improvements in decision-making processes and future action.

#### 3.1.2. Supporting learning and capacity building in QI

Sixteen (16) records were identified as describing learning and training in QI, with a focus on improving capacity among individuals involved in the public health response to COVID-19. The knowledge gained from these activities were typically organizational endeavours that in turn promoted the use of evidence-based QI approaches in public health activities. These included educational resources to support self-directed learning and participation in facilitated activities, such as learning exchanges.^15^ Of note is the importance of harnessing real-time in-practice learning through a reflective approach that was highlighted in two studies. First, Ruebush et al.^18^ described lessons from the implementation of early case investigation and contact-tracing programs from the frontline public health professional perspective, and highlighted future opportunities for this work. They outline several program models and contact tracing collaboratives including standardized training programs, technologies that can improve workflow and community engagement leading to long-term resilience. Additionally, the results of a pandemic exercise by Bacci et al.^11^ highlighted opportunities for public health agencies and their stakeholders to use formal evaluations to build on strengths and improve capacity. Many of the learnings from this exercise became relevant to COVID-19 as participants were able to apply the evaluation findings to establishing testing and vaccination sites throughout Washington State.^11^

Specific grey literature results describe using QI methods and tools to prepare participants for learning, and some were adapted with respect to the management of COVID-19. One example is the Project Public Health Ready program,^19^ which helps local public health agencies build capacity in an intensive 18-month program intended to strengthen partnerships and support the development of an all-hazards response plan in accordance with relevant standards. The First Nations Health Authority (FNHA) also hosts a Community Accreditation and Quality Improvement (CAQI) program to support culturally safe learning and leadership among First Nations communities.^20^ Notably, many of the grey literature resources found in the scoping review did not describe mandated or implemented outcomes, and instead, the level of involvement QI activites was left to the discretion of the participating organizations.

Since the beginning of the COVID-19 pandemic, public health professionals have faced significant demands to increase their capacity and adapt to new challenges to meet the evolving needs of pandemic response. As seen in the literature, improving access to educational resources and building capacity in QI may represent key actions for public health agencies that are interested in developing or bolstering their organizational QI strategies.

#### 3.1.3. Learning from past emergencies

Five (5) records described the use of QI tools and techniques following large-scale health emergencies (e.g., Ebola, Severe acute respiratory syndrome (SARS), H1N1, Zika Virus, etc.). These records described recommendations for improvement from previous emergencies, including measures that could be applied to the COVID-19 pandemic. For instance, the World Health Organization (WHO)^21^ and European Centre for Disease Prevention and Control (ECDC)^22,23^ both encourage the use of IARs and AARs for countries to leverage key opportunities for learning and improvement to better respond to the COVID-19 pandemic. The ECDC^22^ developed a one-day IAR protocol to help countries evaluate lessons learned during COVID-19, with the recognition that abbreviated versions of tools and resources may be valuable during major emergencies where the time and resources to participate in evaluations are limited, in addition to practical constraints (e.g., limited size and frequency of in-person meetings due to physical distancing measures). These activities contribute to organizational QI by empowering stakeholders engaged in the COVID-19 response to identify key strategic issues, challenges, opportunities, and best practices in order to develop relevant and timely solutions that can be readily implemented throughout the ongoing pandemic.^22^

Although the uptake of IARs and AARs by public health agencies has been encouraged by globally prominent public health agencies, an analysis by Parker^24^ found that AARs conducted in the wake of previous health emergencies (e.g., the 2001 Anthrax letter mailings, 2003 SARS epidemic, and others) yielded a pattern of ‘lessons observed but not lessons learned’. This suggests that despite the intention of identifying corrective action to better address future emergencies, these lessons are often neither implemented nor sustained. The author found that despite an increase in interest and resource investments immediately following major infectious disease emergencies, attention typically wanes over subsequent years; thus, highlighting the importance of developing more sustainable policies and funding to support emergency preparedness.^24^ Furthermore, the author indicates that AARs are essential to improving public health emergency preparedness by contributing to the essential evidence-based feedback loop and sustained application within public health practice. A separate study from Marshall et al.,^25^ examining Florida’s response to the 2016-2018 Zika Virus outbreak and assessed collaboration and adaptation across systems of care to provide recommendations for response to future outbreaks, including potential implications for COVID-19. While this study did not conduct an AAR, the WHO Health Systems Framework was used to systematically assess the PHEM response, and utilized journey-mapping and stakeholder engagement techniques to collect data for their evaluation which highlighted several areas of improvement in these indicator areas.^25^

Processes for learning from past events was a key area identified in the literature. This encompassed formal improvement-oriented evaluations, such as IARs and AARs, which help teams and organizations identify critical lessons learned and factors contributing to weaknesses which, if addressed, can improve the response to the next emergency. In the context of infectious disease emergencies, and as observed during the COVID-19 pandemic, the implications are far-reaching and may lead to substantial social, economic and humanitarian consequences on a global scale. As such, learning and improvement in emergency preparedness is crucial for mitigating these harms.

#### 3.1.4. Implementing QI methods during COVID-19

Five (5) records described individual or organizational experiences implementing QI projects or initiatives. The use of QI tools and methods, including process mapping, root cause analysis, and PDSA cycles were found to be effective in supporting the achievement of specific objectives. Aragon et al.^12^ described how local health officers in California applied a variety of frameworks and quality tools to analyze the evolving pandemic situation and facilitate crisis problem-solving. For instance, local health officers used root cause analysis to assess incoming travellers and PDSA cycles to inform decision intelligence. The review also identified recordings describing the use of of QI tools and methods to support the implementation of technology within public health systems, particularly as the COVID-19 pandemic necessitated the reconfiguration of infrastructure and processes in these settings to better support digital and remote work. One examples was an article by Flynn et al.^26^ which discussed the use of computer simulation and process mapping to establish drive-through COVID-19 testing sites. Stakeholders were asked to provid continuous feedback on the drive-through testing program in order to identify and optimize processes relating to traffic flow and staff protocols.^27^ Similarly, Mehta et al.^26^ described the adaptation of Microsoft Teams by a UK National Health Service (NHS) Foundation Trust hospital to improve team communication during the pandemic. Similarly, the NHS developed a Quality Strategy and Quality Framework^28^ including a workbook to help health care agencies evaluate, assess risk and continuously improve health care delivery. The resource was updated to incorporate information from early COVID-19 decision-making.

Despite the substantial strain on public health capacity during COVID-19 pandemic, there is some evidence that the use of established QI tools and techniques guided improvements to PHEM and response work during the COVID-19 pandemic. Although large-scale QI projects may be challenging to plan, execute, and evaluate due to the rapid progression of the COVID-19 pandemic as well as the sizeable human and material resource demands required for response, this scoping review identified several examples of QI applied to generate improvements on a smaller scale (e.g., in teams or individual programs).

#### 3.1.5. Evaluating performance using frameworks/indicators

In health care and public health settings, frameworks and indicators can be useful tools to organize and conceptualize common elements across organizations, and to systematically measure and assess areas for improvement. This review identified seven (7) records that applied a framework or set of established indicators to assess public health management of COVID-19. For example, formal frameworks and indicators were used to assess emergency preparedness and/or response at various stages of the COVID-19 pandemic, as well as the performance of selected countries.

Some studies used established frameworks and indicators as the basis for evaluating the PHEM response to COVID-19. Curtis et al.^29^ applied the US Centers for Disease Control and Prevention (CDC) guidelines to the evaluation of public health surveillance systems to assess the surveillance of COVID-19 patients in Australia. Marshall et al.^25^ applied the WHO Health Systems framework to assess the public health building blocks of health service delivery, health workforce, health information systems, access to essential medicines, financing, and leadership and governance to provide recommendations for response to future outbreaks including COVID-19. This review also found that researchers were interested in assessing public health capabilities at the national level to allow for comparisons across countries and as relevant to a global health context. For instance, Neogi et al.^30^ used the WHO Health System Framework and Global Health Security (GHS) score to assess the health system pandemic preparedness of several countries. Their findings highlighted a notable gap in countries’ health system performance in addressing public health emergencies, regardless of development level.^30^ When compared against their real world responses to COVID-19, GHS scores were not consistent with the results of the Health System Framework in countries such as South Korea, Italy, Spain and Australia.^30^

Kandel et al^31^ similarly used the indicators from the International Health Regulations (IHR) State Party Annual Reporting (SPAR) tool to develop an index that assessed countries’ capacities to prevent, detect, and respond to outbreaks. They found that national capacities varied widely, although there is an overall need to increase the strength of emergency preparedness infrastructure and update national plans.^31^ On the local level, Boyce et al.^32^ proposed a novel framework to rapidly assess urban health security and inform outbreak response efforts. Overall, the use of established frameworks and indicators allowed authors to measure and compare performance and identify strengths and weaknesses within systems at various levels. The information from such assessments are applicable to measure and generate improvements during the COVID-19 pandemic, as well as other health emergencies.

## 4. Discussion

The results from our scoping review illustrate many examples of public health agencies adapting QI and improvement at the project and organizational levels in response to the COVID-19 pandemic. QI concepts, tools and techniques were applied to a variety of PHEM functions and also at different stages of the emergency management cycle, which provides a useful framework for understanding how public health and emergency management systems intersect to respond to infectious health emergencies.^33^ The cycle describes four stages, including preparedness, response, recovery, and mitigation, with each stage representing an action or capacity of PHEM systems to support resilience.^33^ The majority of records described QI activities undertaken prior to the onset of COVID-19, or actions taken during the early stages of the pandemic; often relating to the preparedness and response phases of the emergency management cycle. This might be explained by the time frame during which the scoping review was conducted, and findings may change as scientific investigations and publications related to novel applications of QI during the COVID-19 pandemic continue to expand. Notably, although no records focused exclusively on recovery were found, the recovery period is often viewed as an opportunity for implementing QI to prepare for for future events. In the context of COVID-19, “inter-wave” periods may provide a valuable opportunity to prepare for future waves of the pandemic.

Public health systems involve a large number of stakeholders (e.g., national departments and agencies, laboratories, health care providers, not-for-profit organizations, pharmaceutical manufacturers) and their structure and function can vary widely across jurisdictions. However, it is this scale, complexity and diversity in functions that renders the task of implementing QI practices in public health settings challenging.^4^ Factors such as strong stakeholder engagement at all levels, and communication and coordination across stakeholders to support decision-making are crucial during emergencies requiring complex, multi-sectoral and inter-jurisdictional responses such as the COVID-19 pandemic.^33^ Organization-wide QI activities and the use of QI tools and techniques may assist in strengthening these channels by ensuring PHEM and relevant partners are accustomed to working together and understanding collaboration towards shared goals, as evidenced by one study where public health and pharmacy sector partnerships enhanced COVID-19 vaccination efforts.^11^ Moreover, engaging stakeholders in preliminary preparedness and planning activities promotes streamlining of functions, supports rapid action, and may reduce mistakes or miscommunications during demanding and stressful response periods.

Proponents of QI have noted that to optimize the benefits of QI and elicit large-scale or transformational change, permeation of QI throughout an organization is required. Riley et al.^1^ and Duffy et al.^6^ both describe processes for QI penetration within public health agencies, whereby small QI projects undertaken at the project or unit level (referred to as ‘small qi’) are repeatedly implemented and gradually diffuse into the overarching culture of the organization. Through these small, repeated efforts, QI gradually gains acceptance as an overarching organizational and management philosophy to prioritize continuous measurement of performance towards improvement. The ECDC recommends the use of IARs at least once during an emergency, as participation and dissemination can lead to greater penetration of QI in the organization. However, the absence of ‘top-down’ and ‘bottom-up’ knowledge, competence, and support for QI is a major barrier to widespread implementation within public health systems and their constituent agencies.^1^ As such, it is important for QI to be entrenched in the culture of public health agencies throughout the emergency management cycle and before emergencies occur to ensure optimal response processes. This review identified several studies and practical resources related to QI methods, tools and techniques for training and organizational planning, which can help staff and leadership develop fundamental knowledge and skills. Like IARs, additional QI projects undertaken during COVID-19 are an opportunity for incremental QI permeation within organizations.

Preparedness and learning were common themes from the records reviewed whereby authors assessed lessons learned from the past for the future or otherwise commented on the importance of preparedness for future waves of COVID-19 and future emergencies; thus, representing an iterative and cyclical process for improvement. IARs, AARs, SWOT analyses, and other improvement-related assessments were found to be useful tools for evaluating public health agency responses, including challenges, opportunities, and key learnings. Despite efforts to document experiences and recommendations from previous emergencies, research indicates this information is not drawn on or acted upon in the advent of novel emergencies.^4^ This has been the case for the COVID-19 pandemic, where critics commented on inadequacies in local, national, and global response, despite the availability of pandemic preparedness plans, and abundant guidance written in the wake of large-scale emergencies such as SARS, H1N1, and Ebola.^24,34^ Although the nature, scale, and context of previous emergencies differ from the COVID-19 pandemic, limiting the ability to draw direct comparisons and parlay previous learnings, lessons drawn from previous emergencies are still of exceptional value – but only if these lessons are learned or actioned. Countries that performed relatively well with respect to public health management of COVID-19 effectively adopted lessons learned from past emergencies of a similar nature.^35^

Authors also described their experiences with implementing a variety of QI tools and techniques during COVID-19. Their findings – while contextually and jurisdictionally specific – may offer valuable lessons for others looking to implement similar methods. QI methodologies range from formal QI tools and techniques implemented at the individual team or project level, to frameworks and methods exist for broader implementation of QI throughout the organization. Notably, few of the records found in the scoping review discussed these broader QI frameworks, suggesting that permeation of QI within public health settings is limited or not recorded and disseminated.

Frameworks provide common terminology and an organized way to conceptualize information, and performance indicators provide standardized methods through which to measure performance, allowing comparability over time and across different settings. The availability of performance measurement data is a critical component of improvement in public health. Although the metrics and goals of QI in clinical and health care settings may differ from those in public health, these settings face similar data quality-related challenges. For instance, multiple factors hinder efficient data-sharing, data collection is labour-intensive, and lack of standardization in data collection methods hinders comparison across jurisdictions and over time; these challenges are compounded amid the demands of emergency response, creating a challenge in having reliable data available to inform action.^36^ Our research team developed an evidence-based framework and indicators to conceptualize and measure public health emergency preparedness in Canada, yet similar rigorous frameworks and indicators are limited for other settings (e.g., US).^37,38^ As such, application of appropriate frameworks and indicators is critical in enabling public health agencies to understand their objectives, assess their performance, and provide reliable data to support decision-making throughout the emergency management cycle.

In this review, we found there is applicability and value in implementing QI tools and techniques during rapidly evolving infectious health emergencies, including the COVID-19 pandemic. Yet, the findings highlight a breadth of opportunities for future research and application of QI in PHEM. While the available literature for this review spans a relatively short period of time, as the literature expands post-pandemic, future updates could explore topics emerging from later waves of the pandemic. For instance, none of the records addressed QI tools and techniques applied to health equity initiatives during the COVID-19 pandemic – an important goal within public health and PHEM. Application of QI to such functions (e.g., collection of socio-demographic data during surveillance, equity-informed approaches to testing and vaccine distribution) will help to support equitable public health measures and PHEM strategies given the disproportionate impact of COVID-19 related to population risk and the social determinants of health, as well as low-/middle-income countries, with deleterious impacts on local and global pandemic trajectories.^39,40^

### 4.1. Limitations

This scoping review had a number of limitations. First, due to the rapid nature of the review, additional search methods (e.g., review of reference lists) were not undertaken; therefore, some relevant records may not have been not included. Second, any internal QI initiatives that were not posted publicly (e.g., access to restricted to organizational employees), or available in English language, were not included in the review; thus limiting the findings. Third, there is wide variation in the terminology used to refer to QI and improvement. Although our detailed search strategy sought to include the most commonly used terms, any terminology that does not appear in our search strings was excluded from the findings.

Finally, the information summarized in this review includes records from a limited timeframe of the COVID-19 pandemic. The findings discussed are subject to change as the COVID-19 pandemic progresses, and the corresponding literature evolves and expands.

## 5. Conclusion

Based on the findings of our review, incorporating QI strategies can help public health agencies throughout the emergency management cycle to support key aspects of PHEM and the COVID-19 response. To optimize the benefits of QI methodologies, implementation should occur at the individual project level as well as the widespread integration of QI as part of an organizational management philosophy and culture. The inclusion of QI in public health practice can also provide a systematic and transparent way for public health agencies to monitor progress and improvements in PHEM and in their efforts to meet population health challenges. Future research describing and exploring QI outcome or process measures relevant to public health settings may provide a more in-depth understanding of the mechanisms of organizational change, and may be helpful for informing future PHEM-related policy decisions.

## Data Availability

Data available within the article or its supplementary materials.

## List of Abbreviations

AAR: After-action review
CDC: US Centers for Disease Control and Prevention
ECDC: European Centre for Disease Prevention and Control
GHS: Global Health Security
IAR: In/intra-action review
LHO: Local health officers
NHS: UK National Health Service
OECD: Organisation for Economic Co-operation and Development
PDSA: Plan-Do-Study-Act
PHEM: Public health emergency management
QI: Quality improvement
SWOT: Strengths, weaknesses, opportunities, threats
UK: United Kingdom
US: United States
WHO: World Health Organization

## Declarations

### Ethics approval and consent to participate

Not applicable

### Consent for publication

Not applicable

### Availability of data and materials

The datasets used and/or analysed during the current study are available from the corresponding author on reasonable request.

## Authors’ contributions

CY, MP, and YK were involved in the conceptualization and design of the study. CY and MP were involved in the search, acquisition and management of peer-reviewed and grey literature records. CY, MP, and FG were involved in the literature screening proccess. CY and MP were involved in data extraction and interpretation. CY and MP prepared the initial draft of the manuscript. All authors were involved in review and substantial revisions of the manuscript draft.

## Acknowledgements

The authors would like to thank Public Health Ontario Library staff for their assistance in developing and executing the literature search strategy.

