## Supplemental Appendix 1 for "Rising Through the Pandemic: A scoping review of quality improvement in public health during the COVID-19 pandemic"

### Appendix A. Search Strings

| Database: MEDLINE | | |
|---|---|---|
| **#** | **Searches** | **Results** |
| 1 | COVID-19/ or SARS-CoV-2/ or (Pandemics/ and Coronavirus Infections/) | 50456 |
| 2 | ("COVID-19" or "SARS-CoV-2").nm,os,ox,ps,px,rs,rs. | 40221 |
| 3 | ("2019 corona virus" or "2019 coronavirus" or "2019 ncov" or "corona virus 19" or "corona virus 2019" or "corona virus 2019" or "corona virus disease 19" or "corona virus disease 2019" or "corona virus epidemic*" or "corona virus outbreak*" or "corona virus pandemic*" or "coronavirus 19" or "coronavirus 2019" or "coronavirus 2019" or "coronavirus disease 19" or "coronavirus disease 2019" or "coronavirus epidemic*" or "coronavirus outbreak*" or "coronavirus pandemic*" or "covid 19" or "covid 2019" or "new corona virus" or "new coronavirus" or "novel corona virus" or "novel coronavirus" or "novel human coronavirus" or "sars coronavirus 2" or "sars cov 2" or "sars cov2" or "sars like coronavirus" or "severe acute respiratory syndrome corona virus 2" or "severe acute respiratory syndrome coronavirus 2" or "severe specific contagious pneumonia" or "wuhan corona virus" or "wuhan coronavirus" or 2019ncov or covid19 or covid2019 or ncov or sarscov2 or ((pandemic* or novel or wuhan) adj3 (coronavirus* or "corona virus*" or betacoronavirus* or "beta coronavirus*" or "beta corona virus*" or pneumonia* or SARS or "severe acute respiratory syndrome")) or (pneumonia adj3 (coronavirus* or "corona virus*" or betacoronavirus* or "beta coronavirus*" or "beta corona virus*" or SARS or "severe acute respiratory syndrome")) or "coronavirus response" or "corona virus response").kf,kw,ti. | 88807 |
| 4 | ("2019 corona virus" or "2019 coronavirus" or "2019 ncov" or "corona virus 19" or "corona virus 2019" or "corona virus 2019" or "corona virus disease 19" or "corona virus disease 2019" or "corona virus epidemic*" or "corona virus outbreak*" or "corona virus pandemic*" or "coronavirus 19" or "coronavirus 2019" or "coronavirus 2019" or "coronavirus disease 19" or "coronavirus disease 2019" or "coronavirus epidemic*" or "coronavirus outbreak*" or "coronavirus pandemic*" or "covid 19" or "covid 2019" or "new corona virus" or "new coronavirus" or "novel corona virus" or "novel coronavirus" or "novel human coronavirus" or "sars coronavirus 2" or "sars cov 2" or "sars cov2" or "sars like coronavirus" or "severe acute respiratory syndrome corona virus 2" or "severe acute respiratory syndrome coronavirus 2" or "severe | 17106 |

| | | |
|---|---|---|
| | specific contagious pneumonia" or "wuhan corona virus" or "wuhan coronavirus" or 2019ncov or covid19 or covid2019 or ncov or sarscov2 or ((pandemic* or novel or wuhan) adj3 (coronavirus* or "corona virus*" or betacoronavirus* or "beta coronavirus*" or "beta corona virus*" or pneumonia* or SARS or "severe acute respiratory syndrome")) or (pneumonia adj3 (coronavirus* or "corona virus*" or betacoronavirus* or "beta coronavirus*" or "beta corona virus*" or SARS or "severe acute respiratory syndrome")) or "coronavirus response" or "corona virus response").ab. and (coronavirus* or "corona virus*" or sars* or "severe acute respiratory syndrome").ti. | |
| 5 | ((Influenza, Human/ or Influenza A Virus, H1N1 Subtype/) and Pandemics/) or Middle East Respiratory Syndrome Coronavirus/ or Ebolavirus/ or Hemorrhagic Fever, Ebola/ or SARS Virus/ or Severe Acute Respiratory Syndrome/ or ((Epidemics/ or Pandemics/ or Emergencies/ or Disease Outbreaks/) and (Pneumonia, Viral/ or Respiratory Tract Infections/)) or ((pandemic* or epidemic or "public health emergency of international concern" or PHEIC or ((diseas* or global* or world* or international*) adj2 outbreak*)) and (respiratory or pneumonia)).kf,kw,ti. or ((H1N1 or influenza or "swine flu" or MERS or Ebola).kf,kw,ti. and (pandemic* or outbreak* or epidemic* or "public health emergenc*").ti,ab,kf,kw.) | 79101 |
| 6 | 1 or 2 or 3 or 4 or 5 | 131571 |
| 7 | Quality Improvement/ or Total Quality Management/ or Quality Control/ or *Quality Assurance, Health Care/ or *Quality Indicators, Health Care/ or *"Quality of Health Care"/ or Qualitative Research/ or Organizational Innovation/ or Efficiency, Organizational/ or *Efficiency/ or Capacity Building/ or "Delivery of Health Care"/st, og or *Outcome Assessment, Health Care/ or *"Outcome and Process Assessment, Health Care"/ or *"Process Assessment (Health Care)"/ or Health Care Evaluation Mechanisms/ or Health care surveys/ or Medical audit/ or Needs Assessment/ or "Health Services Needs and Demand"/og, st or Health Care Reform/ or Health Services Accessibility/st, og or Health Services Administration/ or Practice Guidelines as Topic/st or Health Planning Guidelines/ or Health Systems Plans/st or Health Plan Implementation/st or Health Planning/st or Regional Health Planning/st or State Health Plans/st or Planning Techniques/ or Program Development/ or Program Evaluation/ or Evaluation Studies as Topic/ or Evaluation Study/ or Health Services Research/ or Operations Research/ or Organizational Case Studies/ or "Time and Motion Studies"/ or Systems Analysis/ or "Task Performance and Analysis"/ or Benchmarking/ or Emergency Medical Services/st or Disaster Planning/st or Public | 954565 |

| | | |
|---|---|---|
| | Health Administration/st or Education, Public Health Professional/ or Competency-Based Education/ or Public Health Practice/st or Public Health/st | |
| 8 | (quality adj3 (improv* or strengthen* or manag* or control* or measur* or indicator* or metric* or assur* or evaluat* or apprais* or audit* or assess* or standard* or benchmark*)).ti,kf,kw. or ("CQI" or "continuous quality improv*" or "quality improv*" or "QI" or "LEAN" or "six sigma" or "intra action report*" or "IAR" or "Strengths, Weaknesses, Opportunities, and Threats" or "SWOT").ti,ab,kf,kw. or (((perform* or performance) adj2 (measur* or indicator* or metric* or evaluat* or apprais* or audit* or assess* or standard* or benchmark* or quality)) or (program* adj2 (evaluat* or develop*)) or "organi?ation* innovat*" or "organi?ation* standard*" or "lessons learned" or "future directions").ti,kf,kw. | 199258 |
| 9 | 7 or 8 | 1087766 |
| 10 | Public Health/ or Public Health Practice/ or Public Health Administration/ or world health organization/ or pan american health organization/ or "united states dept. of health and human services"/ or united states public health service/ or "centers for disease control and prevention, u.s."/ or "national institutes of health (u.s.)"/ or "united states agency for healthcare research and quality"/ or "united states health resources and services administration"/ or "national health planning information center, u.s."/ or (("public health" adj2 (agency or agencies or organi?ation* or institut* or system* or practic* or administration)) or ((center* or centre*) adj2 "disease control") or "world health organization" or "cent* for disease control" or "national institutes of health" or "department of health and human services" or "public health ontario" or "health canada" or "public health agency of canada" or "BCCDC" or "INSPQ").ti,kf,kw. or ("public health".ti,kf,kw. not medline.st.) | 192993 |
| 11 | (government agencies/ or government programs/ or government/ or exp health planning organizations/ or exp "state health planning and development agencies"/ or international agencies/ or International Cooperation/ or red cross/ or International Health Regulations/ or Global Health/ or local government/ or state government/) and "public health".ti,kf,kw. | 6381 |
| 12 | 10 or 11 | 195258 |
| 13 | 6 and 9 and 12 | 606 |
| 14 | ((*Disaster Planning/ or *Disasters/pc or *Emergencies/ or *Disease Outbreaks/pc or *Epidemics/pc or *Pandemics/pc or *Civil Defense/ or Mass Casualty Incidents/ or Medical Countermeasures/ or Strategic Stockpile/ or Surge Capacity/) and (*Public Health/ or *Public Health Practice/ or *Public Health Administration/)) or | 2636 |

| | | |
|---|---|---|
| | (((((emergency or emergencies or pandemic* or epidemic* or outbreak*) adj2 (manag* or mitigat* or prepar* or plan or plans or plann* or respon* or ready or readiness or recover* or countermeasure* or operation* or system* or "public health")) or "rapid respons*" or "crisis manag*" or "crisis communicat*" or "system* preparedness" or "communit* preparedness" or "contingenc* plan*" or "International Preparedness & Response to Emergencies & Disasters" or "IPRED").ti,kf,kw. not medline.st.) and "public health".ti,kf,kw.) or ("public health emergency manag*" or "PHEM" or "public health emergency prepar*" or "public health emergency respons*").ti,kf,kw. | |
| 15 | 9 and 14 | 599 |
| 16 | 13 or 15 | 1088 |
| 17 | (exp Africa/ or exp Caribbean Region/ or exp Central America/ or exp Latin America/ or exp South America/ or exp Asia/ or Developing Countries/) not (north america/ or exp canada/ or mexico/ or exp united states/ or chile/ or israel/ or turkey/ or exp japan/ or exp "republic of korea"/ or europe/ or austria/ or belgium/ or baltic states/ or estonia/ or latvia/ or lithuania/ or czech republic/ or hungary/ or poland/ or slovakia/ or slovenia/ or exp france/ or exp germany/ or exp united kingdom/ or greece/ or ireland/ or exp italy/ or luxembourg/ or netherlands/ or portugal/ or "scandinavian and nordic countries"/ or denmark/ or greenland/ or finland/ or iceland/ or norway/ or svalbard/ or sweden/ or spain/ or switzerland/ or exp australia/ or new zealand/ or exp Developed Countries/) | 1026461 |
| 18 | 16 not 17 | 890 |
| 19 | 18 not (hospital or hospitals or hospital-based or clinic or clinics or clinician* or "emergency room*" or "emergency department*" or "emergency ward*" or "emergency care" or "emergency medic*" or surger* or surgical* or "intensive care" or "ICU").ti. | 858 |
| 20 | limit 19 to yr="2003 -Current" | 818 |
| 21 | limit 20 to english | 797 |

| **Database: Ovid EMBASE (Search 1 of 2)** | | | |
|---|---|---|---|
| **#** | | **Searches** | **Results** |
| 1 | | coronavirus disease 2019/ or severe acute respiratory syndrome coronavirus 2/ | 80443 |
| 2 | | pandemic/ and Coronavirus infection/ | 10224 |

| | | | |
|---|---|---|---|
| 3 | | ("2019 corona virus" or "2019 coronavirus" or "2019 ncov" or "corona virus 19" or "corona virus 2019" or "corona virus 2019" or "corona virus disease 19" or "corona virus disease 2019" or "corona virus epidemic*" or "corona virus outbreak*" or "corona virus pandemic*" or "coronavirus 19" or "coronavirus 2019" or "coronavirus 2019" or "coronavirus disease 19" or "coronavirus disease 2019" or "coronavirus epidemic*" or "coronavirus outbreak*" or "coronavirus pandemic*" or "covid 19" or "covid 2019" or "new corona virus" or "new coronavirus" or "novel corona virus" or "novel coronavirus" or "novel human coronavirus" or "sars coronavirus 2" or "sars cov 2" or "sars cov2" or "sars like coronavirus" or "severe acute respiratory syndrome corona virus 2" or "severe acute respiratory syndrome coronavirus 2" or "severe specific contagious pneumonia" or "wuhan corona virus" or "wuhan coronavirus" or 2019ncov or covid19 or covid2019 or ncov or sarscov2 or ((pandemic* or novel or wuhan) adj3 (coronavirus* or "corona virus*" or betacoronavirus* or "beta coronavirus*" or "beta corona virus*" or pneumonia* or SARS or "severe acute respiratory syndrome")) or (pneumonia adj3 (coronavirus* or "corona virus*" or betacoronavirus* or "beta coronavirus*" or "beta corona virus*" or SARS or "severe acute respiratory syndrome")) or "coronavirus response" or "corona virus response").af. | 98415 |
| 4 | | pandemic influenza/ or 2009 H1N1 influenza/ or "Influenza A virus (H1N1)"/ or ebola hemorrhagic fever/ or ebolavirus/ or zaire ebolavirus/ or severe acute respiratory syndrome/ or sars-related coronavirus/ or middle east respiratory syndrome/ or middle east respiratory syndrome coronavirus/ | 32982 |
| 5 | | (emergency/ or epidemic/ or pandemic/) and (virus pneumonia/ or respiratory tract infection/) | 12718 |
| 6 | | ((pandemic* or epidemic* or "public health emergency of international concern" or PHEIC or ((diseas* or global* or world* or international*) adj2 outbreak*)) and (respiratory or pneumonia)).kw,ti. or ((H1N1 or influenza or "swine flu" or MERS or Ebola).kw,ti. and (pandemic* or outbreak* or epidemic* or "public health emergenc*").ti,kw.) | 16036 |
| 7 | | 1 or 2 or 3 or 4 or 5 or 6 | 135232 |
| 8 | | "program cost effectiveness"/ or "public reporting (health care)"/ or "utilization review"/ or benchmarking/ or clinical audit/ or evaluation study/ or good laboratory practice/ or *health care planning/ or health care quality/ or health care survey/ or *health services research/ or *needs assessment/ or organizational development/ or organizational efficiency/ or performance measurement system/ | 939404 |

| | | | |
|---|---|---|---|
| | | or professional standard/ or program acceptability/ or program appropriateness/ or program development/ or program effectiveness/ or program efficacy/ or program evaluation/ or program feasibility/ or program impact/ or program sustainability/ or public health systems research/ or quality control/ or *planning techniques/ or system analysis/ or task performance/ or total quality management/ or *capacity building/ or *"organization and management"/ | |
| 9 | | (disaster planning/ or emergency health service/ or health care access/ or health care availability/ or health care distribution/ or health care planning/ or health care system/ or public health/ or public health service/) and standard/ | 19222 |
| 10 | | (quality adj3 (improv* or strengthen* or manag* or control* or measur* or indicator* or metric* or assur* or evaluat* or apprais* or audit* or assess* or standard* or benchmark*)).ti,kw. or ("CQI" or "continuous quality improv*" or "quality improv*" or "QI" or "LEAN" or "six sigma" or "intra action report*" or "IAR" or "Strengths, Weaknesses, Opportunities, and Threats" or "SWOT").ti,ab,kw. or (((perform* or performance) adj2 (measur* or indicator* or metric* or evaluat* or apprais* or audit* or assess* or standard* or benchmark* or quality)) or (program* adj2 (evaluat* or develop*)) or "organi?ation* innovat*" or "organi?ation* standard*" or "lessons learned" or "future directions").ti,kw. | 278695 |
| 11 | | 8 or 9 or 10 | 1112879 |
| 12 | | *public health/ or public health service/ or european medicines agency/ or health canada/ or "international federation of red cross and red crescent societies"/ or "medicines and healthcare products regulatory agency"/ or national health service/ or world health organization/ | 300774 |
| 13 | | (("public health" adj2 (agency or agencies or organi?ation* or institut* or system* or practic* or administration)) or ((center* or centre*) adj2 "disease control") or "world health organization" or "cent* for disease control" or "national institutes of health" or "department of health and human services" or "public health ontario" or "health canada" or "public health agency of canada" or "BCCDC" or "INSPQ").ti,kw. or ("public health".ti,kw. not embase.st.) | 47947 |
| 14 | | government/ and international cooperation/ and "public health".ti,kw. | 68 |
| 15 | | 12 or 13 or 14 | 322464 |
| 16 | | 7 and 11 and 15 | 820 |
| 17 | | 16 not (hospital or hospitals or hospital-based or clinic or clinics or clinician* or "emergency room*" or "emergency department*" or "emergency ward*" or | 781 |

| | | | |
|---|---|---|---|
| | | "emergency care" or "emergency medic*" or surger* or surgical* or "intensive care" or "ICU").ti. | |
| 18 | | (exp Africa/ or exp Asia/ or exp "South and Central America"/ or exp Eastern Europe/ or developing country/) not (exp North America/ or Chile/ or exp "Australia and New Zealand"/ or Europe/ or exp Western Europe/ or exp Southern Europe/ or Japan/ or korea/ or south korea/ or czech republic/ or hungary/ or poland/ or slovakia/ or slovenia/ or israel/ or "turkey (republic)"/) | 1355702 |
| 19 | | 17 not 18 | 611 |
| 20 | | limit 19 to conference abstracts | 14 |
| 21 | | 19 not 20 | 597 |
| 22 | | 21 not (editorial or letter or note).pt. | 402 |
| 23 | | limit 22 to yr="2003 -Current" | 400 |
| 24 | | limit 23 to english language | 379 |

| Database: Ovid EMBASE (Search 2 of 2) | | |
|---|---|---|
| **#** | **Searches** | **Results** |
| 1 | (disaster planning/ or disaster/pc or *emergency/ or epidemic/pc or pandemic/pc or civil defense/ or mass disaster/ or medical countermeasure/ or surge capacity/ or relief work/) and (public health/ or public health service/) | 3801 |
| 2 | (((((emergency or emergencies or pandemic* or epidemic* or outbreak*) adj2 (manag* or mitigat* or prepar* or plan or plans or plann* or respon* or ready or readiness or recover* or countermeasure* or operation* or system* or "public health")) or "rapid respons*" or "crisis manag*" or "crisis communicat*" or "system* preparedness" or "communit* preparedness" or "contingenc* plan*" or "International Preparedness & Response to Emergencies & Disasters" or "IPRED").ti,kw. not embase.st.) and "public health".ti,kw.) or ("public health emergency manag*" or "PHEM" or "public health emergency prepar*" or "public health emergency respons*").ti,kw. | 892 |
| 3 | 1 or 2 | 4314 |
| 4 | "program cost effectiveness"/ or "utilization review"/ or benchmarking/ or evaluation study/ or health care quality/ or health care survey/ or *health services research/ or *needs assessment/ or organizational development/ or organizational efficiency/ or performance measurement system/ or professional standard/ or program acceptability/ or program appropriateness/ or program development/ or program effectiveness/ or program efficacy/ or program evaluation/ or program feasibility/ or | 1593683 |

| | | |
|---|---|---|
| | program impact/ or program sustainability/ or public health systems research/ or quality control/ or *planning techniques/ or system analysis/ or task performance/ or total quality management/ or *capacity building/ or *"organization and management"/ or qualitative research/ or outcome assessment/ or *professional competence/ or *practice guideline/ or *standard/ | |
| 5 | (quality adj3 (improv* or strengthen* or manag* or control* or measur* or indicator* or metric* or assur* or evaluat* or apprais* or audit* or assess* or standard* or benchmark*)).ti,kw. or ("CQI" or "continuous quality improv*" or "quality improv*" or "QI" or "LEAN" or "six sigma" or "intra action report*" or "IAR" or "Strengths, Weaknesses, Opportunities, and Threats" or "SWOT").ti,ab,kw. or (((perform* or performance) adj2 (measur* or indicator* or metric* or evaluat* or apprais* or audit* or assess* or standard* or benchmark* or quality)) or (program* adj2 (evaluat* or develop*)) or "organi?ation* innovat*" or "organi?ation* standard*" or "lessons learned" or "future directions").ti,kw. | 280311 |
| 6 | 4 or 5 | 1749781 |
| 7 | 3 and 6 | 849 |
| 8 | 7 not (hospital or hospitals or hospital-based or clinic or clinics or clinician* or "emergency room*" or "emergency department*" or "emergency ward*" or "emergency care" or "emergency medic*" or surger* or surgical* or "intensive care" or "ICU").ti. | 802 |
| 9 | (exp Africa/ or exp Asia/ or exp "South and Central America"/ or exp Eastern Europe/ or developing country/) not (exp North America/ or Chile/ or exp "Australia and New Zealand"/ or Europe/ or exp Western Europe/ or exp Southern Europe/ or Japan/ or korea/ or south korea/ or czech republic/ or hungary/ or poland/ or slovakia/ or slovenia/ or israel/ or "turkey (republic)"/) | 1363421 |
| 10 | 8 not 9 | 691 |
| 11 | limit 10 to conference abstracts | 9 |
| 12 | 10 not 11 | 682 |
| 13 | 12 not (editorial or letter or note).pt. | 603 |
| 14 | limit 13 to yr="2003 -Current" | 574 |
| 15 | limit 14 to english language | 559 |

| **Database: Ovid Global Health (Search 1 of 2)** | | |
|---|---|---|
| **#** | **Searches** | **Results** |

| | | |
|---|---|---|
| 1 | ((((emergency or emergencies or pandemic* or epidemic* or outbreak*) adj2 (manag* or mitigat* or prepar* or plan or plans or plann* or respon* or ready or readiness or recover* or countermeasure* or operation* or system* or "public health")) or "rapid respons*" or "crisis manag*" or "crisis communicat*" or "system* preparedness" or "communit* preparedness" or "contingenc* plan*" or "International Preparedness & Response to Emergencies & Disasters" or "IPRED") and "public health").ti,sh,id. or ("public health emergency manag*" or "PHEM" or "public health emergency prepar*" or "public health emergency respons*").ti,ab,id,sh. | 11059 |
| 2 | (quality adj3 (improv* or strengthen* or manag* or control* or measur* or indicator* or metric* or assur* or evaluat* or apprais* or audit* or assess* or standard* or benchmark*)).ti,sh,id. or ("CQI" or "continuous quality improv*" or "quality improv*" or "QI" or "LEAN" or "six sigma" or "intra action report*" or "IAR" or "Strengths, Weaknesses, Opportunities, and Threats" or "SWOT").ti,ab,id,sh. or (((perform* or performance) adj2 (measur* or indicator* or metric* or evaluat* or apprais* or audit* or assess* or standard* or benchmark* or quality)) or (program* adj2 (evaluat* or develop*)) or "organi?ation* innovat*" or "organi?ation* standard*" or "lessons learned" or "future directions").ti,sh,id. | 47016 |
| 3 | 1 and 2 | 192 |
| 4 | 3 not (hospital or hospitals or hospital-based or clinic or clinics or clinician* or "emergency room*" or "emergency department*" or "emergency ward*" or "emergency care" or "emergency medic*" or surger* or surgical* or "intensive care" or "ICU").ti. | 187 |
| 5 | limit 4 to yr="2003 -Current" | 187 |
| 6 | limit 5 to english language | 182 |

| **Database: Ovid Global Health (Search 2 of 2)** | | |
|---|---|---|
| **#** | **Searches** | **Results** |
| 1 | ("2019 corona virus" or "2019 coronavirus" or "2019 ncov" or "corona virus 19" or "corona virus 2019" or "corona virus 2019" or "corona virus disease 19" or "corona virus disease 2019" or "corona virus epidemic*" or "corona virus outbreak*" or "corona virus pandemic*" or "coronavirus 19" or "coronavirus 2019" or "coronavirus 2019" or "coronavirus disease 19" or "coronavirus disease 2019" or "coronavirus epidemic*" or "coronavirus outbreak*" or "coronavirus pandemic*" or "covid 19" or "covid 2019" or "new corona virus" or "new coronavirus" or "novel corona virus" or "novel coronavirus" or "novel human coronavirus" or "sars coronavirus 2" or "sars cov | 16563 |

| | | |
|---|---|---|
| | 2" or "sars cov2" or "sars like coronavirus" or "severe acute respiratory syndrome corona virus 2" or "severe acute respiratory syndrome coronavirus 2" or "severe specific contagious pneumonia" or "wuhan corona virus" or "wuhan coronavirus" or 2019ncov or covid19 or covid2019 or ncov or sarscov2 or ((pandemic* or novel or wuhan) adj3 (coronavirus* or "corona virus*" or betacoronavirus* or "beta coronavirus*" or "beta corona virus*" or pneumonia* or SARS or "severe acute respiratory syndrome")) or (pneumonia adj3 (coronavirus* or "corona virus*" or betacoronavirus* or "beta coronavirus*" or "beta corona virus*" or SARS or "severe acute respiratory syndrome")) or "coronavirus response" or "corona virus response").id,sh,ti. | |
| 2 | ("2019 corona virus" or "2019 coronavirus" or "2019 ncov" or "corona virus 19" or "corona virus 2019" or "corona virus 2019" or "corona virus disease 19" or "corona virus disease 2019" or "corona virus epidemic*" or "corona virus outbreak*" or "corona virus pandemic*" or "coronavirus 19" or "coronavirus 2019" or "coronavirus 2019" or "coronavirus disease 19" or "coronavirus disease 2019" or "coronavirus epidemic*" or "coronavirus outbreak*" or "coronavirus pandemic*" or "covid 19" or "covid 2019" or "new corona virus" or "new coronavirus" or "novel corona virus" or "novel coronavirus" or "novel human coronavirus" or "sars coronavirus 2" or "sars cov 2" or "sars cov2" or "sars like coronavirus" or "severe acute respiratory syndrome corona virus 2" or "severe acute respiratory syndrome coronavirus 2" or "severe specific contagious pneumonia" or "wuhan corona virus" or "wuhan coronavirus" or 2019ncov or covid19 or covid2019 or ncov or sarscov2 or ((pandemic* or novel or wuhan) adj3 (coronavirus* or "corona virus*" or betacoronavirus* or "beta coronavirus*" or "beta corona virus*" or pneumonia* or SARS or "severe acute respiratory syndrome")) or (pneumonia adj3 (coronavirus* or "corona virus*" or betacoronavirus* or "beta coronavirus*" or "beta corona virus*" or SARS or "severe acute respiratory syndrome")) or "coronavirus response" or "corona virus response").ab. and (coronavirus* or "corona virus*" or sars* or "severe acute respiratory syndrome").ti. | 5374 |
| 3 | ((pandemic* or epidemic or "public health emergency of international concern" or PHEIC or ((diseas* or global* or world* or international*) adj2 outbreak*)) and (respiratory or pneumonia)).id,sh,ti. or ((H1N1 or influenza or "swine flu" or MERS or Ebola).id,sh,ti. and (pandemic* or outbreak* or epidemic* or "public health emergenc*").ti,ab,id,sh.) | 17364 |
| 4 | 1 or 2 or 3 | 33103 |

| | | |
|---|---|---|
| 5 | (quality adj3 (improv* or strengthen* or manag* or control* or measur* or indicator* or metric* or assur* or evaluat* or apprais* or audit* or assess* or standard* or benchmark*)).ti,sh,id. or ("CQI" or "continuous quality improv*" or "quality improv*" or "QI" or "LEAN" or "six sigma" or "intra action report*" or "IAR" or "Strengths, Weaknesses, Opportunities, and Threats" or "SWOT").ti,ab,id,sh. or (((perform* or performance) adj2 (measur* or indicator* or metric* or evaluat* or apprais* or audit* or assess* or standard* or benchmark* or quality)) or (program* adj2 (evaluat* or develop*)) or "organi?ation* innovat*" or "organi?ation* standard*" or "lessons learned" or "future directions").ti,sh,id. | 46825 |
| 6 | (("public health" adj2 (agency or agencies or organi?ation* or institut* or system* or practic* or administration)) or ((center* or centre*) adj2 "disease control") or "world health organization" or "cent* for disease control" or "national institutes of health" or "department of health and human services" or "public health ontario" or "health canada" or "public health agency of canada" or "BCCDC" or "INSPQ").ti,sh,id. | 13319 |
| 7 | public health/ or WHO/ or ((public agencies/ or international organizations/) and "public health".ti,sh,id.) | 133836 |
| 8 | 6 or 7 | 135039 |
| 9 | 4 and 5 and 8 | 73 |
| 10 | 9 not (hospital or hospitals or hospital-based or clinic or clinics or clinician* or "emergency room*" or "emergency department*" or "emergency ward*" or "emergency care" or "emergency medic*" or surger* or surgical* or "intensive care" or "ICU").ti. | 72 |
| 11 | limit 10 to yr="2003 -Current" | 72 |
| 12 | limit 11 to english language | 71 |

| **Database: Scopus (Search 1 of 2)** | | |
|---|---|---|
| **#** | **Queries** | **Results** |
| 1 | ( TITLE-ABS-KEY ( ( ( ( emergency OR emergencies OR pandemic* OR epidemic* OR outbreak* ) W/2 ( manag* OR mitigat* OR prepar* OR plan OR plans OR plann* OR respon* OR ready OR readiness OR recover* OR countermeasure* OR operation* OR system* OR "public health" ) ) OR "rapid respons*" OR "crisis manag*" OR "crisis communicat*" OR "system* preparedness" OR "communit* preparedness" OR "contingenc* plan*" OR "International Preparedness & Response to Emergencies & Disasters" OR "IPRED" ) AND "public health" ) ) OR ( TITLE-ABS-KEY ( "public health emergency manag*" OR "PHEM" OR "public health emergency prepar*" OR "public health emergency respons*" ) ) | 13,461 |

| | | |
|---|---|---|
| 2 | TITLE-ABS-KEY ( quality W/3 ( improv* OR strengthen* OR manag* OR control* OR measur* OR indicator* OR metric* OR assur* OR evaluat* OR apprais* OR audit* OR assess* OR standard* OR benchmark* ) ) OR TITLE-ABS-KEY ( "CQI" OR "continuous quality improv*" OR "quality improv*" OR "QI" OR "LEAN" OR "six sigma" OR "intra action report*" OR "IAR" OR "Strengths, Weaknesses, Opportunities, and Threats" OR "SWOT" ) OR TITLE-ABS-KEY ( ( ( perform* OR performance ) W/2 ( measur* OR indicator* OR metric* OR evaluat* OR apprais* OR audit* OR assess* OR standard* OR benchmark* OR quality ) ) OR ( program* W/2 ( evaluat* OR develop* ) ) OR "organi?ation* innovat*" OR "organi?ation* standard*" OR "lessons learned" OR "future directions" ) | 2,971,608 |
| 3 | #1 AND #2 | 1,549 |
| 4 | #3 NOT TITLE ( hospital OR hospitals OR hospital-based OR clinic OR clinics OR clinician* OR "emergency room*" OR "emergency department*" OR "emergency ward*" OR "emergency care" OR "emergency medic*" OR surger* OR surgical* OR "intensive care" OR "ICU" ) | 1,393 |
| 5 | #4 AND LANGUAGE ( english ) AND PUBYEAR > 2002 | 1,289 |
| 6 | #5 AND NOT ( INDEX ( medline ) ) | 304 |

| **Database: Scopus (Search 2 of 2)** | | |
|---|---|---|
| **#** | **Queries** | **Results** |
| 1 | TITLE-ABS-KEY ( ( "2019 corona virus" OR "2019 coronavirus" OR "2019 ncov" OR "corona virus 19" OR "corona virus 2019" OR "corona virus 2019" OR "corona virus disease 19" OR "corona virus disease 2019" OR "corona virus epidemic*" OR "corona virus outbreak*" OR "corona virus pandemic*" OR "coronavirus 19" OR "coronavirus 2019" OR "coronavirus 2019" OR "coronavirus disease 19" OR "coronavirus disease 2019" OR "coronavirus epidemic*" OR "coronavirus outbreak*" OR "coronavirus pandemic*" OR "covid 19" OR "covid 2019" OR "new corona virus" OR "new coronavirus" OR "novel corona virus" OR "novel coronavirus" OR "novel human coronavirus" OR "sars coronavirus 2" OR "sars cov 2" OR "sars cov2" OR "sars like coronavirus" OR "severe acute respiratory syndrome corona virus 2" OR "severe acute respiratory syndrome coronavirus 2" OR "severe specific contagious pneumonia" OR "wuhan corona virus" OR "wuhan coronavirus" OR 2019ncov OR covid19 OR covid2019 OR ncov OR sarscov2 OR ( ( pandemic* OR novel OR wuhan ) W/3 ( coronavirus* OR "corona virus*" OR betacoronavirus* OR "beta coronavirus*" OR "beta corona virus*" OR pneumonia* OR sars OR "severe acute respiratory syndrome" ) ) OR ( pneumonia W/3 ( coronavirus* OR "corona virus*" OR betacoronavirus* OR "beta coronavirus*" OR "beta corona virus*" OR sars OR "severe acute respiratory syndrome" ) ) OR "coronavirus response" OR "corona virus response" ) ) | 107,930 |
| 2 | TITLE-ABS-KEY ( ( ( pandemic* OR epidemic OR "public health emergency of international concern" OR pheic OR ( ( diseas* OR global* OR world* OR international* ) W/2 outbreak* ) ) AND ( respiratory OR pneumonia ) ) ) | 71,300 |
| 3 | TITLE-ABS-KEY ( h1n1 OR influenza OR "swine flu" OR mers OR ebola ) AND TITLE-ABS-KEY ( pandemic* OR outbreak* OR epidemic* OR "public health emergenc*" ) | 50,217 |
| 4 | #1 OR #2 OR #3 | 167,191 |

| | | |
|---|---|---|
| 5 | TITLE-ABS-KEY ( quality W/3 ( improv* OR strengthen* OR manag* OR control* OR measur* OR indicator* OR metric* OR assur* OR evaluat* OR apprais* OR audit* OR assess* OR standard* OR benchmark* ) ) OR TITLE-ABS-KEY ( "CQI" OR "continuous quality improv*" OR "quality improv*" OR "QI" OR "LEAN" OR "six sigma" OR "intra action report*" OR "IAR" OR "Strengths, Weaknesses, Opportunities, and Threats" OR "SWOT" ) OR TITLE-ABS-KEY ( ( ( perform* OR performance ) W/2 ( measur* OR indicator* OR metric* OR evaluat* OR apprais* OR audit* OR assess* OR standard* OR benchmark* OR quality ) ) OR ( program* W/2 ( evaluat* OR develop* ) ) OR "organi?ation* innovat*" OR "organi?ation* standard*" OR "lessons learned" OR "future directions" ) | 2,960,087 |
| 6 | TITLE-ABS-KEY ( ( "public health" W/2 ( agency OR agencies OR organi?ation* OR institut* OR system* OR practic* OR administration ) ) OR ( ( center* OR centre* ) W/2 "disease control" ) OR "world health organization" OR "cent* for disease control" OR "national institutes of health" OR "department of health and human services" OR "public health ontario" OR "health canada" OR "public health agency of canada" OR "BCCDC" OR "INSPQ" ) | 266,357 |
| 7 | #4 AND #5 AND #6 | 707 |
| 8 | #7 NOT TITLE ( hospital OR hospitals OR hospital-based OR clinic OR clinics OR clinician* OR "emergency room*" OR "emergency department*" OR "emergency ward*" OR "emergency care" OR "emergency medic*" OR surger* OR surgical* OR "intensive care" OR "ICU" ) | 662 |
| 9 | #8 AND LANGUAGE ( english ) AND PUBYEAR > 2002 | 629 |
| 10 | #9 AND NOT ( INDEX ( medline ) ) | 146 |
